# Perceived Implications of Current Conflict of Interest Policy in Individuals Accepted or Rejected for Participation in Food and Drug Administration Advisory Committee

**DOI:** 10.1101/2020.11.25.20237438

**Authors:** Yuichi J. Shimada, Kyle Hair, Adrienne Faerber, Christian G. Zimmerman, Timothy Houchin, Robert A. Hart, Matthew D. Gibb, Shunichi Homma

**Author notes:** **Address for correspondence:** Dr. Shunichi Homma, MD, MHCDS, Columbia University Medical Center, 177 Ft. Washington Avenue, Milstein Hospital Building, 5th Floor, 5GN, Room 435, New York, NY, 10032.

## Abstract

**Background and Objective:** The Food and Drug Administration (FDA) relies on advice from scientific and medical experts to make approval decisions about new prescription medications and medical devices. Therefore, it is crucial that FDA Advisory Committees (ACs) include the most knowledgeable scientists and clinicians in the decision-making process. However, to ensure that the advice is free from biases, current FDA policy often excludes those most qualified from participating in ACs due to perceived conflicts of interest (COI). The objective of the present study is to elicit opinion among subject matter experts regarding current FDA COI rules and regulations.

**Methods:** We conducted a cross-sectional study using formal, self-administered online survey consisting of 14 questions, 3 of which addressed perceived implications of current FDA COI policy. We send a formal online survey to study subjects via Qualtrics. Study subjects were 1) individuals who participated in FDA ACs and 2) those who were interested in participating in ACs and had completed FDA COI paperwork but rejected by the FDA due to COI. The outcome measure is response to the 3 questions about the perceived implications of current FDA COI policy. Responses were scored on a 5-point Likert-type scale.

**Results:** Among 403 study subjects (200 AC members and 203 individuals who were rejected due to COI), 145 (36%) responded to the survey including 90 AC members and 55 rejected individuals. Respondents included 41 (28%) females. 97% were holding MD or PhD degrees. 88% were age 46 and over, and 66% had more than 25 years in practice. The primary findings were that 49% of respondents agreed that the current FDA regulations lead to a lower quality of experts on ACs, 72% agreed that current policies exclude qualified experts from serving on ACs, and 48% agreed that FDA policy lowers the overall quality of AC input, to at least a “moderate” extent (19%-37% to a “high” or “very high” extent).

**Conclusions:** The prevailing opinion among respondents to the formal survey is that current FDA COI policy has the potential effects to lowering quality of experts, excluding qualified experts, and lowering overall quality of AC input. The present report elucidates a potential need for the FDA to discuss the benefit and risk of the current AC COI policies.

## INTRODUCTION

The US Food and Drug Administration (FDA) oversees more than $2.5 trillion in drugs, devices, and other consumables.^1^ Millions of Americans depend on the FDA to make approval decisions for the novel drugs, biologics, and devices that come to market each year.^1^ During the decision-making process, the FDA frequently relies on Advisory Committees (ACs) to provide expert scientific, technical, and policy advice, and the FDA typically follows the recommendations of ACs.^2, 3^ Therefore, ACs have come under increased scrutiny in recent years, specifically with respect to members’ financial and professional relationships with regulated industries.^4,5^

ACs consist of expert scientists and practitioners from diverse healthcare fields who serve as special government employees on 50 standing panels that evaluate drugs, biotechnology and vaccines, and medical devices.^6^ More than 900 expert scientists and clinicians serve as special government employees on ACs and are subject to current conflicts of interest (COI) regulations.^7^ Frequently, the experts needed to provide sound advice to the federal government are also in high demand by the industries being regulated, namely drug and device makers. This so-called ‘shared pool dilemma,’ where the most highly qualified experts sought as consultants by the FDA are those most likely to have financial ties to regulated industries, has been the subject of vigorous debate among scientists, policy makers, and even within the FDA itself.^8^ Numerous op-eds have appeared in the scientific literature and mainstream newspapers, and high-profile scandals in the early 2000s brought the shared-pool dilemma into the public spotlight.^4^ One effect of the shared pool dilemma has been increased regulations aimed at minimizing potential COI among AC members.

Notwithstanding the legitimate need to address potential conflicts of interest in AC members, there is also a growing consensus among stakeholders that it would be extremely difficult – although perhaps not impossible – to replace current levels of AC expertise with non-conflicted panels.^9, 10^ Many current and former FDA employees and expert consultants feel that selection policies and processes for ACs must evolve to maintain the optimum levels of expertise needed by the FDA in its decision-making process. However, there has been no formal survey research conducted to gauge the collective opinion among current subject matter experts.

Therefore, we sought to perform a pilot study to evaluate the extent to which expert scientists and clinicians believe current FDA regulations result in a lower quality of expert consultants, exclude qualified experts, and lower the overall quality of AC input.

## METHODS

### Study subjects

Study subjects were all individuals who were interested in participating in FDA ACs and had completed FDA COI paperwork from May 1, 2016 to April 30, 2017. Study subjects were stratified based on the outcome of the COI process. FDA provided a list of individuals who had no COI and were allowed to participate in an FDA AC (N=200) and those who were interested in participating in an FDA AC and completed the COI paperwork but rejected due to potential COI (N=203). All 403 individuals on the list were invited to respond to the survey.

### Survey

A survey was created using Qualtrics (www.qualtrics.com) and consisted of 14 questions. Three questions specifically addressed perceived implications of current FDA COI policy as follows:

1. To what extent, if any, does current FDA COI policy lead to a lower quality of expert consultants?
2. To what extent, if any, does current FDA COI policy exclude qualified experts to serve on FDA drug or device ACs?
3. To what extent, if any, does current FDA COI policy lower the overall quality of AC input?

The rest of the questions addressed basic demographics and previous inclusion/exclusion in FDA ACs. Participants answered questions on a 5-point Likert-type scale. For questions 1-3, the responses were “to a very high level,” “to a high level,” “to a moderate level,” “to a low level,” and “to no level.” The final survey question was a free-text box for participants to provide any additional comments or thoughts on the FDA COI policies and the AC selection process. The survey was sent to participants through an email invitation on August 9, 2017, and participants received up to four reminders to complete the survey. The survey was closed on September 28, 2017.

Responses were collected and analyzed via Qualtrics. Missing data was handled in a pairwise fashion: if a participant answered at least one question, their results were included in the analysis. Thus, respondents did not need to complete the entire survey for their responses to be included in the analysis. Free-text responses were grouped according to similar themes and based on positive or negative tone of the response.

### Statistical analysis

Statistical comparisons between AC members versus individuals who were rejected due to COI were conducted using Chi-Square analysis, using Bonferroni method to control for multiple comparisons (statistical significance threshold for p < 0.007).

## RESULTS

Of the 403 study subjects receiving the survey – i.e., 200 AC members and 203 individuals who were rejected due to COI, 145 (36%) responded to at least one question. Of 145 who responded to at least one question, the response rate for individual questions ranged from 71% to 100%. **Table 1** presents the demographic and experience characteristics of survey respondents. Respondents were stratified into two groups – AC members (N=90) versus individuals who were rejected due to COI (N=55). The AC members and rejected individuals were similar in age, gender, terminal degree, years of experience, and participation in drug versus device committees. Overall, self-reported level of familiarity with FDA ACs was “very high” or “high” in 74% of respondents, while 26% reported either “moderate” or “a little” level of familiarity. Of those participants who had previously been rejected from serving on an AC, 68% responded that they were given a reason for their rejection. Of those, 61% of participants served on drug committees only, with the remaining participants serving on device (23%) or both drug and device (16%) committees.

**Table 1.** Baseline characteristics of the survey participants according to Advisory Committee membership

|  | **Advisory**<br>**Committee**<br>**Members**<br>**(N = 90)** | **Rejected due to**<br>**Conflict of**<br>**Interest**<br>**(N = 55)** | ***P* value** |
| --- | --- | --- | --- |
| Age, years (%) |  |  |  |
| 25 to 35 | 2 (2.4) | 0 | .39 |
| 36 to 45 | 11 (12.9) | 4 (7.3) |  |
| 46 to 60 | 28 (32.9) | 23 (41.8) |  |
| 61 or older | 44 (51.8) | 28 (50.9) |  |
| Gender (%) |  |  |  |
| Male | 54 (62.8) | 45 (83.3) | .02 |
| Female | 32 (37.2) | 9 (16.7) |  |
| Degree (%) |  |  |  |
| MD | 53 (72.6) | 38 (71.7) | .81 |
| PhD | 17 (23.3) | 14 (26.4) |  |
| Other (APRN, JD) | 3 (4.1) | 1 (1.9) |  |
| Experience in practice, years (%) |  |  |  |
| 0 to 9 | 2 (2.6) | 0 | .40 |
| 10 to 25 | 24 (30.8) | 20 (37.0) |  |
| 26 or more | 52 (66.7) | 34 (63.0) |  |
| Committee Participation (%) |  |  |  |
| Drug | 59 (80.8) | 19 (61.3) | .06 |
| Device | 14 (19.2) | 12 (38.7) |  |

Data were expressed as numbers (percentages).

Response rates for the three questions addressing perceived implications of FDA COI policy ranged from 81% to 85%. Results from these questions are shown in **Figure 1**. Among the respondents, 58 (49%) reported that current FDA COI policy lowers the quality of experts to a “moderate”, “high”, or “very high” extent (**Figure 1**, Panel A). Similarly, 89 (72%) indicated that FDA COI policy excludes qualified experts and 56 (48%) answered that FDA COI policy lowers the overall quality of ACs to at least “moderate” extent (**Figure 1**, Panel B, C). To these questions, 19%-37% of respondents answered “high” or “very high” (**Figure 1**).

**Figure 1.**
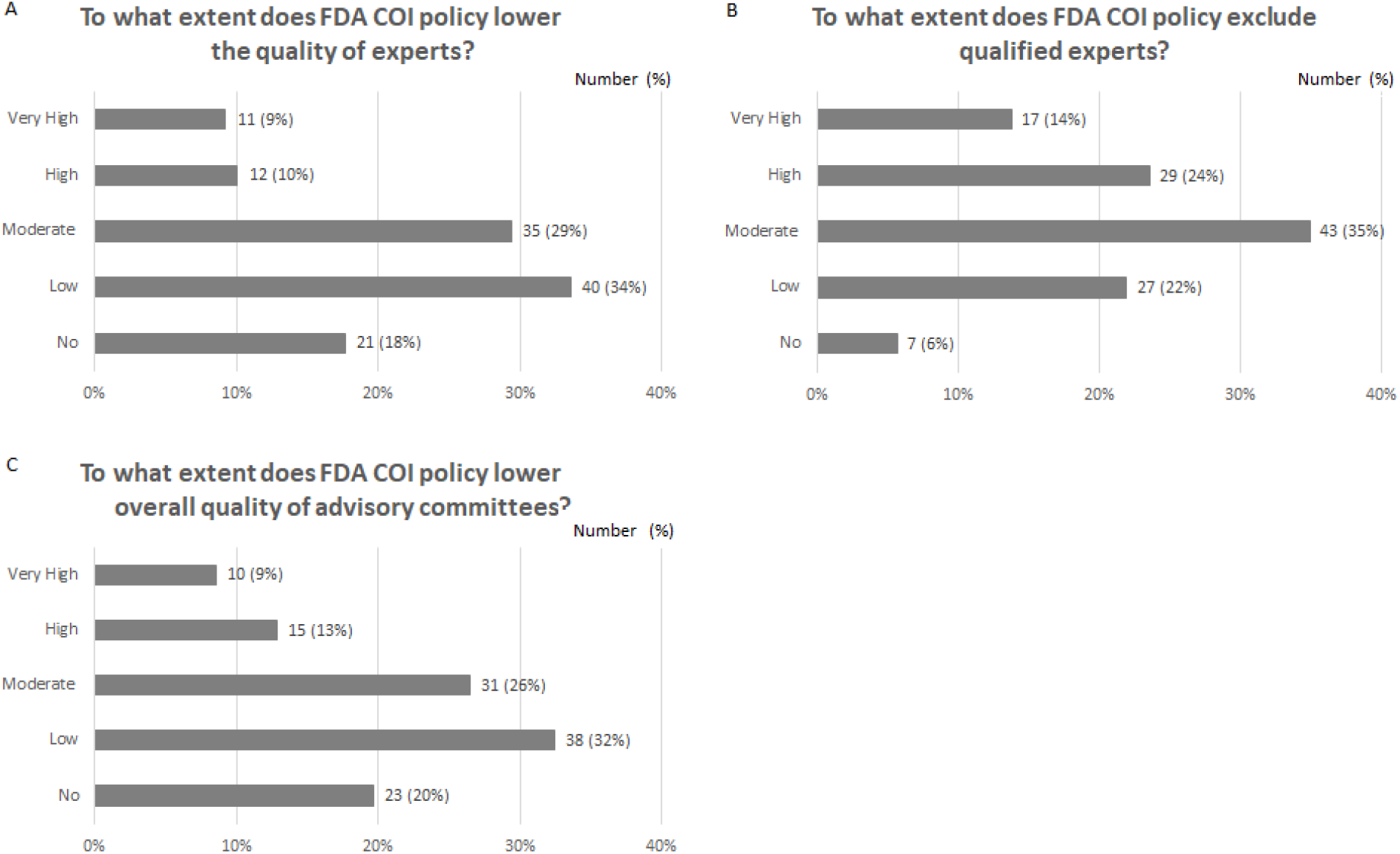
Perceived implications of current FDA COI policy on AC process. Study subjects were asked to state to what extent current COI policy lowers quality of experts (panel A), excludes qualified experts (panel B), and lowers the overall quality of the AC input (panel C). Each bar represents number of study subjects responding in each category. Abbreviations: COI, conflict of interest; AC, advisory committee.

Comparisons between AC members versus individuals who were rejected due to COI (**Figure 2**) revealed that AC members were more likely to report lower extent to which FDA COI lowers quality of experts (*P* < .001), excludes qualified experts (*P* < .001), and lowers the quality of AC input (*P* = .004).

**Figure 2.**
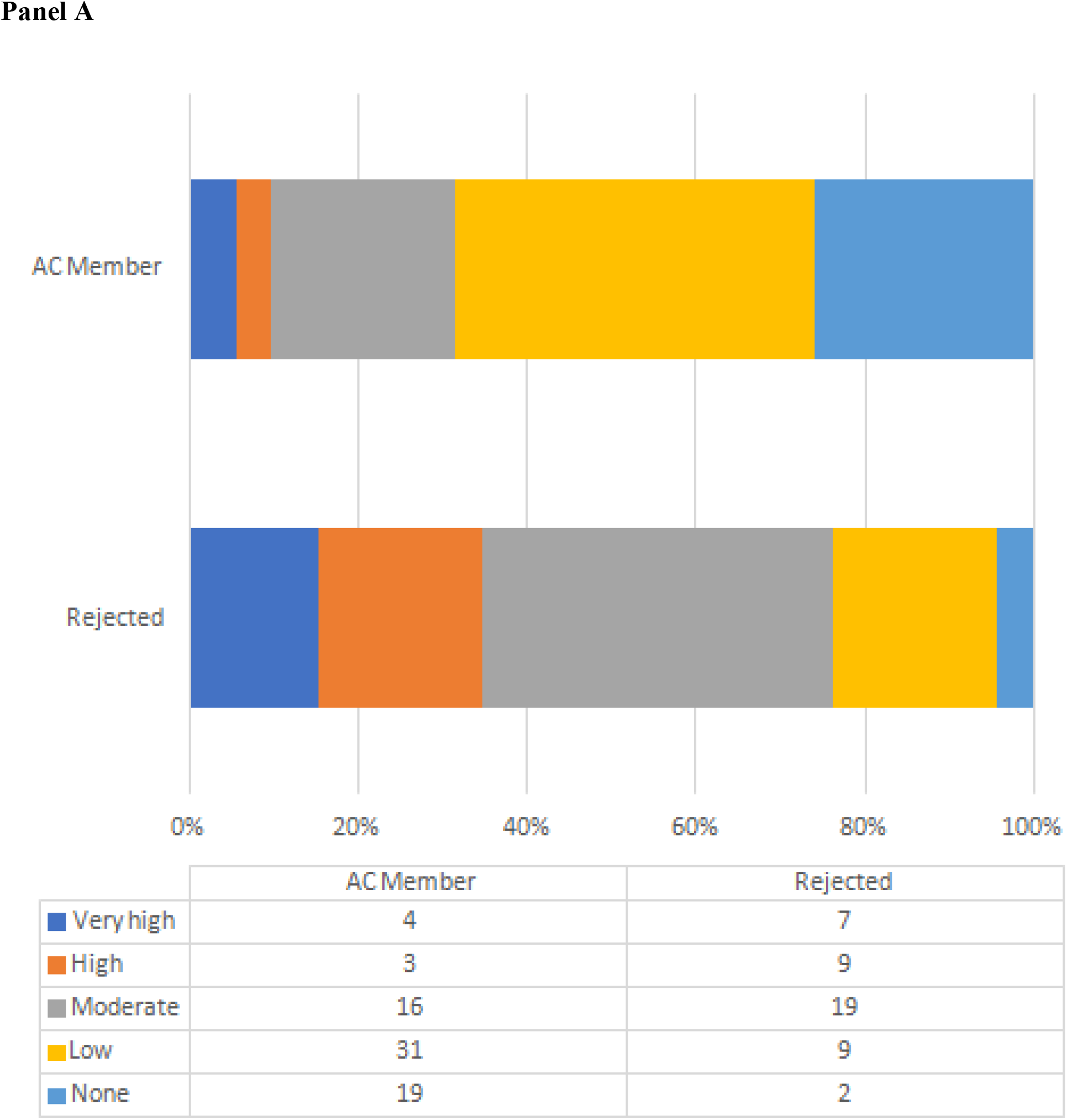
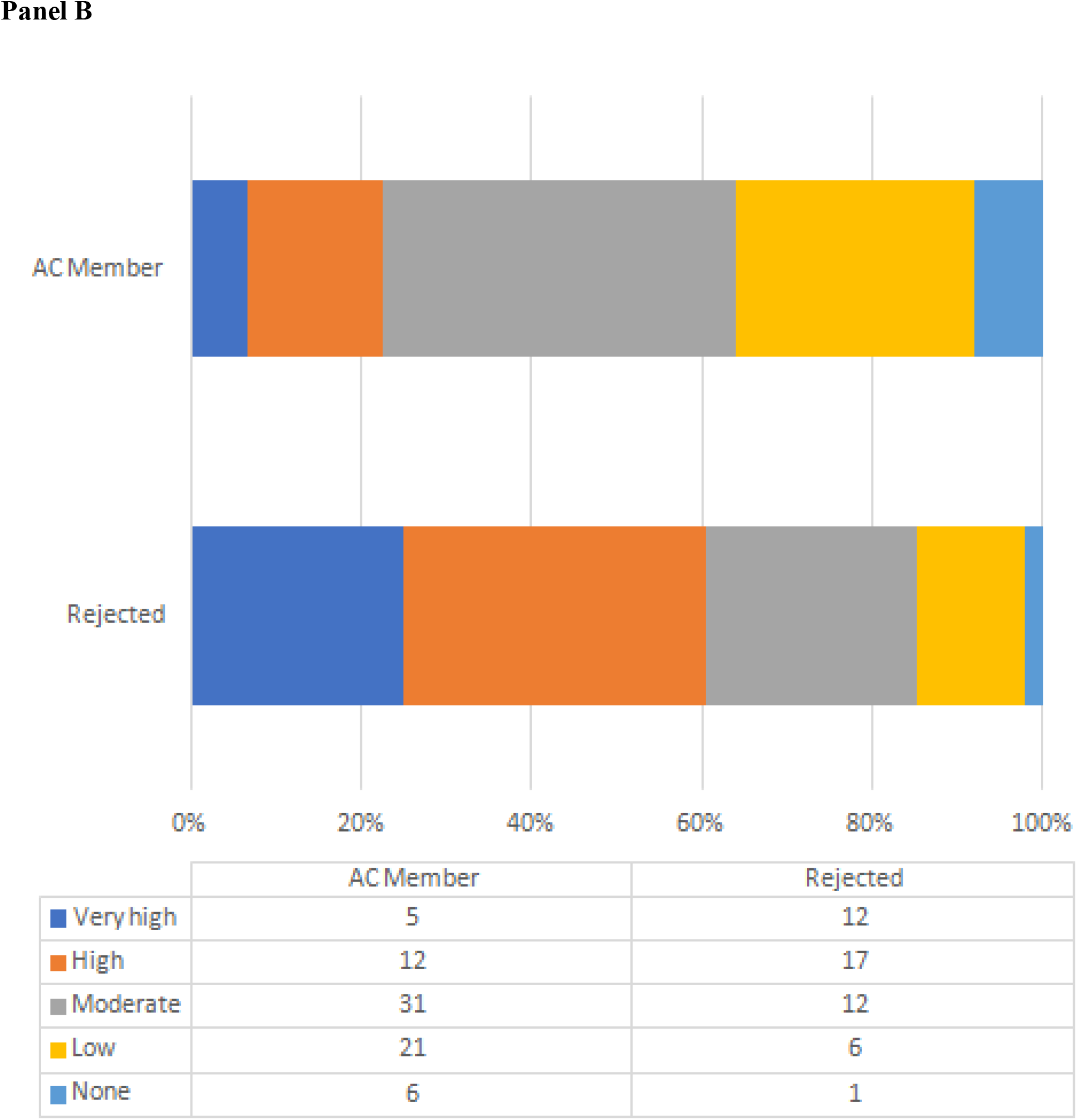
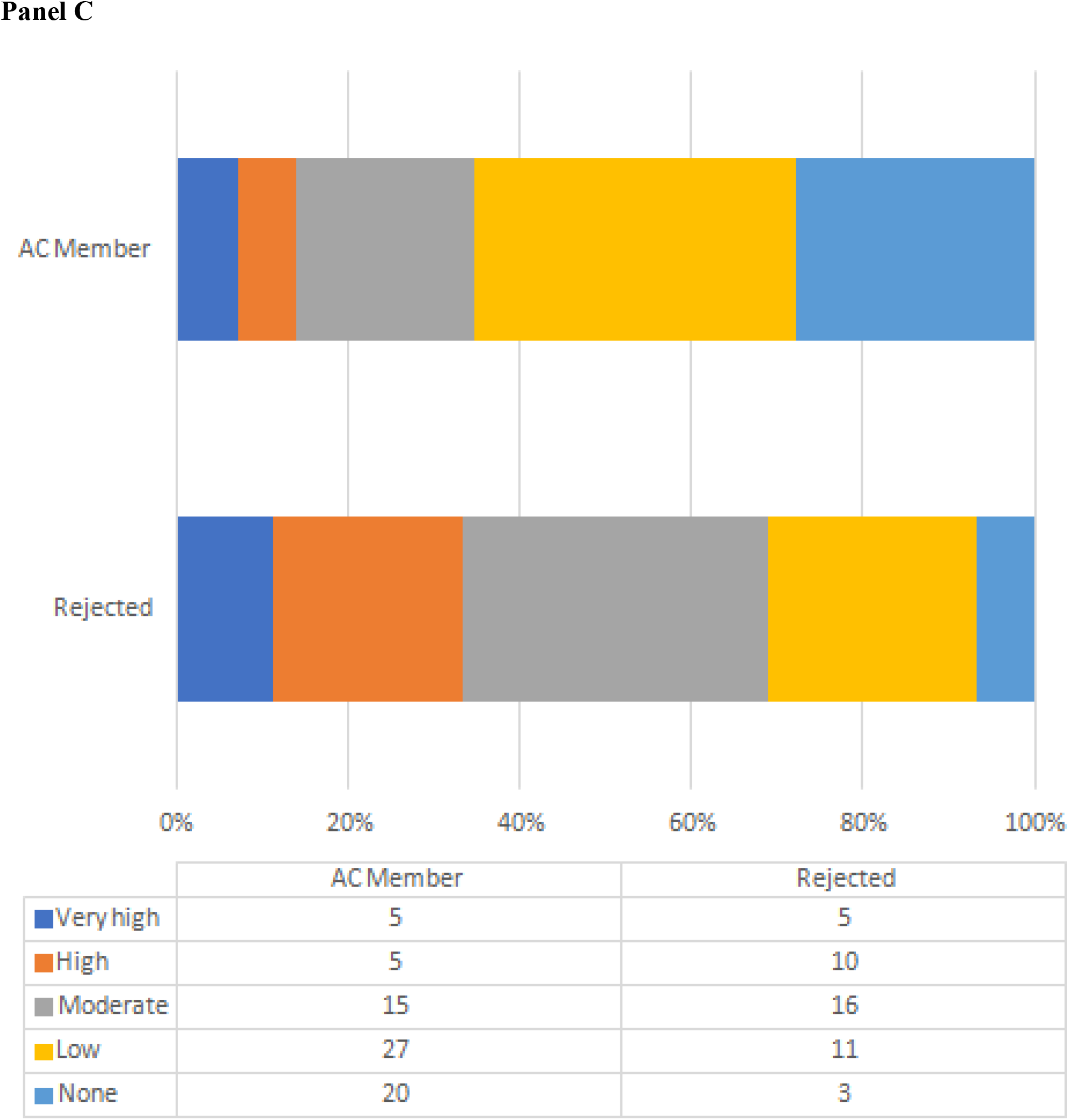
Perceived implications of current FDA COI policy on AC process, stratified by AC membership. Study subjects were asked to state to what extent current COI policy lowers quality of experts (panel A), excludes qualified experts (panel B), and lowers the overall quality of the AC input (panel C). Abbreviations: COI, conflict of interest; AC, advisory committee

Nearly one-third of respondents provided free-text feedback regarding current COI regulations. Feedback from participants is summarized into the following themes: 1) There were both positive and negative attitudes and beliefs about the FDA COI process. 2) Respondents who reported being rejected from ACs were more negative in their attitudes and beliefs about the FDA COI process. 3) There was frustration regarding what constituted a financial COI, including blanket financial exclusions and COIs arising from professional affiliations. 4) Dissatisfaction was expressed with regard to the process (e.g., time, paperwork) involved in determining whether a COI exists.

## DISCUSSION

In this formal opinion survey from subject matter experts with a high degree of familiarity with the COI process including both individuals who were accepted and rejected to participate in AC, more than a half responded that current FDA policy leads to a lower quality of expert consultants, excludes relevant experts, and reduces the overall quality of the AC process to at least a moderate degree. The results of this study add to the body of literature on FDA COI policy by providing important insights into the perceived impact of current FDA COI policies by the subject matter experts.

### Results in context

Current or potential AC members who have a real or imputed COI must disclose the information to the FDA prior to serving as a voting member on an AC. Such conflicts include current investments, employment, consulting and speaking arrangements, contracts, grants, and patents. Spousal conflicts of interest must also be disclosed. In addition, there is an imputed COI based on financial holdings with one’s employer or other affiliated organization; individuals with no personal financial holdings in a regulated company may be considered conflicted under these current ‘blanket’ rules and get excluded from participating in ACs.

Eliminating all financial relationships might not be feasible but it is crucial to keep the financial conflicts at a low level in committee meetings; otherwise ties to pharmaceutical companies can introduce undesired influence on members’ voting.^11, 12^ Because of increasingly complex legislation, the FDA is often unable to fill panels with non-conflicted experts.^13^ At its discretion, and following the requirements under U.S.C. 208(b), the FDA may grant waivers to individuals with disclosed COIs to bring essential expertise into the AC.^14^ Although the majority of waivers are given, the process for granting waivers has become more complex over time, and many experts in academia and within the FDA feel that current regulations have resulted in a failure to obtain the optimum level of expertise needed in ACs (U.S. Food and Drug Administration, 2017). ^15^

In accordance with these prior reports, half of respondents in the present study supported the idea that current regulations lower the overall quality of experts on AC panels, with 49% of respondents agreeing, to at least moderate extent, that the quality of experts may be lowered. Furthermore, many respondents agreed that current regulations exclude qualified experts from participating in ACs (72% agreement) and lower the overall quality of AC input (48% agreement). These findings are in line with other studies, as well as the opinion of current and former FDA officials, that current policy likely excludes qualified experts.^15, 16^

Another interesting observation in the current study was the different responses between the AC members and individuals who had been rejected participation due to COI. The AC members more frequently answered that current FDA COI policy compromises some of the quality measures of ACs (i.e., quality of experts, quality of AC input). On the other hand, they were more likely to report lower extent to which the current FDA COI policy excludes qualified experts. These findings indicate that the perceived effects of current FDA COI policy on the quality of ACs may be different according to prior rejection due to potential COI.

### Impact of COI on AC input

Although the law includes stipulations to reduce the effect of member COIs,^17^ only a handful of studies have examined the relationship between financial COIs and AC voting behavior at the meeting level. A widely-cited study in 2006 observed that removal of all conflicted AC members would not have changed the outcome in any of their decisions.^18^ The empirical evidence supports essentially no association between meeting-level conflicts and FDA approval decisions.^16, 19^ Meanwhile, there is a growing consensus among stakeholders that current policies limiting conflicted members from participating in ACs reduce the competence of FDA decision-making and poses an increased cost to Americans who rely on the FDA to make sound decisions.^16, 19^ In two separate FDA-commissioned studies in 2013 and 2015, researchers found that there was no significant evidence that AC member votes were compromised by their financial interest.^16, 19^ In their 2013 report, they further stated that the ability to create conflict-free ACs “would represent an uncertain and potentially substantial additional burden on the cost and the timeliness of AC operations”.^16^ The 2015 study examined AC meetings for drugs between 1997 and 2012, and no evidence was found that conflicted ACs were more likely to vote for drug approval than non-conflicted panels.^19^ Moreover, conflicted members’ votes were more often in line with the FDA’s ultimate decision when compared with non-conflicted members.

The report concluded that “if anything…conflicts are associated with a lower propensity to recommend drug approval, which is the opposite of the concern that has motivated recent reforms”.^19^ These reports indicate that removal of all voting members with sponsor conflicts would not result in any change in approval decisions by the FDA, with a caveat that the Searle Civil Justice Institute was funded by the Searle Freedom Trust, which was founded by a pharmaceutical executive.

There are two possible extreme outcomes of the shared pool dilemma – one is to have the most qualified experts on ACs with some of them necessarily having COIs, and the other is to exclude conflicted experts and have less qualified AC members to decide the fate of a drug or device than their conflicted counterparts.^20^ The prior reports and inferences from the present study would call for further discussion of the ongoing FDA COI policy to improve the quality of AC members, include qualified experts, and eventually augment quality of AC input. Based on the free-text feedback in the current survey, we propose that a few examples of potential modifications would be 1) elimination of blanket exclusion of universities and other organizations that have benefited from industry grants, 2) exemption of mutual funds and certain institutional investments from COI (e.g., 401K, 403b, and pension accounts), and 3) increasing the current threshold for individual holding (i.e., US$50,000 per year).

### Limitations

The present pilot study has several limitations. First, only less than half of the experts who were invited to participate in the survey responded, which may introduce biases (e.g., selection bias and non-response bias). For example, some studies have demonstrated that multiple variables such as age, gender, location, and socioeconomic status differ between respondents and non-respondents to self-answered surveys.^21, 22^ These unmeasured and measured differences in baseline characteristics between respondents and non-respondents might have skewed the survey results. Second, the current survey did not examine the association between COI and the individual or AC level input, as this was above the scope of the current analysis. Third, this was a cross-sectional study and does not provide information to infer causality. Fourth, perceived benefits of the current FDA COI policy were not included in the questionnaire. Last, the current study did not address the burden (e.g., time, paperwork) associated with the COI determination process.

## Conclusions

The present pilot surveillance study, involving FDA AC members and those who had been rejected due to potential COI, elucidated that more than a half of these experts perceive current FDA COI policy as reducing the quality of AC members, rejecting qualified experts, and resulting in lower quality of AC input. The present study would further facilitate the conduct of a larger, more comprehensive survey to examine how the current FDA policy is viewed, which would provide an insightful input on how the FDA policy could be more balanced between benefit and harm to the quality of AC.

## Data Availability

The data that support the findings of this study are available on request from the corresponding author. The data are not publicly available.

## DECLARATION OF INTEREST STATEMENT

### Conflict of Interest Disclosures

Dr. Houchin is a Behavioral Health Medical Director for WellCare Health Plans. Dr. Hart has the following disclosures: Honoraria from Globus and DepuySynthes, Royalties from Seaspine and DupuySynthes, Study Funding for Other Projects from ISSGF, Speaking and/or Teaching Arrangements from DepuySynthes, Board membership from CSRS (Scientific Advisory Board), ISSLS (Scientific Advisory Board), ISSG (Scientific Advisory Board), and AOSpine (Technical Committee), and Grants from Medtronic (past) and ISSG (current). The other authors have no disclosures.

### Funding/Support

None

## Author Contributions

Dr. Homma had full access to all of the data in the study and take responsibility for the integrity of the data and the accuracy of the data analysis.

*Study concept and design:* Hair, Gibb, and Homma.

*Acquisition of data:* Hair, Faerber, Gibb, and Homma.

*Analysis and interpretation of data:* Hair, Faerber, Zimmerman, Houchin, Hart, Gibb, and Homma.

*Drafting of the manuscript:* Shimada.

*Critical revision of the manuscript for important intellectual content:* Hair, Faerber, Zimmerman, Houchin, Hart, Gibb, and Homma.

*Statistical expertise:* Shimada and Faerber.

*Study supervision:* Gibb and Homma.

## Acknowledgements

The authors would like to thank Mark Whiting for assistance with preparing the written manuscript.

## Notes

### Author Declarations

The Columbia University Institutional Review Boards (Columbia University Administrative Review Committee Protocol Number: IRB-AAAS6808) determined that this study qualified as exempt, and no consent form was required.

